# Prevalence and Multi-Domain Determinants of Elevated Blood Lead Levels Among Pregnant Women in Bangladesh: Evidence from the 2025 Multiple Indicator Cluster Survey

**DOI:** 10.64898/2026.10.12.26365283

**Authors:** Syed Jayedul Bashar, Md Fuad Al Fidah, Md. Ridwan Islam, Anjan Kumar Roy, Rubhana Raqib, Tahmeed Ahmed, Mustafa Mahfuz

## Abstract

**Background:** Lead is a cumulative environmental toxicant for which no safe level of exposure has been identified, and pregnant women and their fetuses are particularly vulnerable because lead readily crosses the placenta and can be mobilised from maternal bone stores. Bangladesh conducted its first nationally representative blood lead testing among pregnant women through the 2025 Multiple Indicator Cluster Survey (MICS), but a detailed, multi-domain analysis of the determinants of elevated blood lead level (BLL) among this population has not yet been reported.

**Methods:** We conducted a secondary cross-sectional analysis of 1,928 pregnant women aged 15– 49 years with complete data from the women’s dataset of Bangladesh MICS 2025. Elevated BLL was defined as ≥5 μg/dL. Survey-weighted logistic regression was used to estimate unadjusted and adjusted odds ratios (aOR) for co-exposure to arsenic, cadmium and mercury and sociodemographic and socioeconomic covariates. Slope Index of Inequality (SII) as well as Relative Index of Inequality (RII) were used to quantify wealth-related inequality. Population attributable fractions (PAF) were estimated for three counterfactual scenarios, and adaptive least absolute shrinkage and selection operator (LASSO) regression was used as a sensitivity analysis.

**Results:** Elevated BLL was present among 6.2% of participants. In the adjusted model, a 2-fold rise in blood cadmium was associated with higher odds of elevated BLL (aOR: 1.54; 95% CI: 1.21–1.96), whereas a 2-fold rise in blood mercury (aOR: 0.63; 95% CI: 0.48–0.83), rural residence (aOR: 0.55; 95% CI: 0.32–0.94), secondary education (aOR: 0.49; 95% CI: 0.28–0.85) and post-secondary education (aOR: 0.22; 95% CI: 0.11–0.46) were associated with lower odds. Belonging to progressively wealthier quintiles was associated with markedly higher odds, BLL rising to an aOR of 10.32 (95% CI: 4.41–24.15) for the richest versus poorest quintile. The RII was 8.59 (95% CI: 4.24–17.40) and the SII was 0.159 (95% CI: 0.101–0.218), indicating a substantial pro-rich gradient in elevated BLL. Counterfactual scenarios suggested that shifting the population to secondary-level education could reduce the prevalence of elevated BLL by 52.9% (95% CI: 20.6–72.0) and shifting to rural residence by 24.3% (95% CI: 1.4–41.9). Adaptive LASSO variable selection produced estimates consistent in direction and magnitude with the fully adjusted model.

**Conclusion:** In this national sample, approximately one out of every 16 pregnant women had blood lead levels that met or exceeded the action threshold set by the World Health Organization, with this issue being particularly prevalent among wealthier, urban, and less-educated women, and was compounded by co-exposure to cadmium. In Bangladesh, the typical pattern observed for maternal and child health indicators does not align with the inverse relationship found between household wealth and lead exposure. This suggests that the sources of exposure are not merely indicators of material poverty. Antenatal blood lead screening, source investigation, and equity-informed lead-reduction strategies that specifically reach wealthier and urban households are warranted.

## Introduction

Lead is a naturally occurring but non-essential heavy metal with no biological function and no threshold below which, it can be considered safe for human health (1). Despite the near-universal phase-out of leaded petrol, global exposure remains widespread: modelling based on the Global Burden of Disease study estimated that lead exposure caused more than 5.5 million premature adult deaths from cardiovascular disease and a loss of over 765 million IQ points in children in 2019, at an economic cost comparable to that of ambient fine particulate air pollution (2). A joint UNICEF–Pure Earth analysis estimated that around one in three children worldwide, nearly 800 million children, have a blood lead level (BLL) at or above 5 μg/dL. Close to half of these children live in South Asia (3). The WHO’s 2021 guideline for the clinical management of lead exposure formally adopted 5 μg/dL as the trigger for exposure investigation while emphasizing that no BLL can be considered free of risk (4). The CDC has established a population-based reference value for blood lead levels in children at 3.5 μg/dL. (5).

Pregnant mothers and their fetus constitute a notably susceptible group. Lead is able to pass through the placenta via passive diffusion and has been identified in fetal brain tissue as soon as it approaches to the ending of the first trimester (6). As bone is the primary reservoir for more than 90% of the adult body burden of lead (7), during pregnancy, the body’s natural rise in bone turnover, prompted by the fetus’s need for calcium, can release lead that has built up in a woman’s body over her lifetime into her bloodstream. Thus, pregnant woman’s BLL corresponds to both of her current environmental exposure and years of prior accumulation (8). Clinical trials suggest that calcium supplementation usually lowers maternal BLL, presumably via suppressing this bone resorption, support the biological plausibility of this pathway (9). Elevated maternal BLL has been associated with a number of unfavorable conditions like gestational hypertension, spontaneous abortion, reduced fetal growth and low birth weight, and impaired offspring neurodevelopment, prompting professional bodies such as the American College of Obstetricians and Gynecologists to recommend targeted screening and counselling once BLL reaches 5 μg/dL (6, 10).

Lead exposure rarely occurs alone. Cadmium, arsenic and mercury frequently cooccur with lead through shared dietary, occupational and environmental pathways, including contaminated food, cookware, cosmetics and biomass or tobacco smoke (11, 12). A recent study concluded that both cadmium and lead are associated with lower birth weight and higher risk of pre-eclampsia, while mercury exposure has been associated with impaired fetal growth, underscoring the importance of examining these metals together rather than lead in isolation (13).

In Bangladesh, data on pregnant women exposed to lead had, until recently, come only from a few, geographically limited studies. A case-control study of 430 pregnant women in three rural districts reported that 31% of the participants had a BLL exceeding 5 μg/dL and indicated lead-containing food cans, agrochemicals and adulterated turmeric as sources (14). Bangladesh has separately been identified as one of the countries most affected by childhood lead poisoning globally, due to informal and unsafe recycling of lead-acid batteries repeatedly flagged as a leading contamination pathway (15). In 2025, Bangladesh conducted its very first nationally representative assessment of blood lead, cadmium, arsenic and mercury in the country’s history as part of round seven of the MICS program, testing pregnant women aged 15–49 years and children aged 12–59 months. The preliminary report identified that 38.3% of children and 7.5% of pregnant women had an elevated BLL, with the highest burden concentrated in Dhaka division and, notably, among the richest wealth quintile rather than the poorest, a pattern that runs reverse to the socioeconomic gradient commonly observed for maternal and child health indicators in this survey series (16).

Albeit this preliminary national report was descriptive, it did not incluide a formal multivariable analysis of the determinants of elevated BLL particularly among pregnant women. In this group, exposure reflects both ongoing environmental contact and the cumulative skeletal lead burden mobilized for the developing fetus. A more comprehensive analysis that integrates co-exposure to other toxic metals, sociodemographic and socioeconomic covariates, formal quantification of wealth-related inequality, and estimation of the population-level impact of plausible policy scenarios would help determine which women are most affected and which policy levers might most effectively reduce this burden. We therefore used data from the Bangladesh MICS 2025 to estimate the national prevalence of elevated BLL among pregnant women and to examine its multi-domain determinants, including co-exposure to arsenic, cadmium and mercury, and sociodemographic and socioeconomic factors, while additionally quantifying wealth-related inequality using the Slope and Relative Indices of Inequality and estimating the population attributable fraction of elevated BLL under selected counterfactual scenarios.

## Methods

### Study design and data source

This is a secondary cross-sectional analysis of data from the Bangladesh Multiple Indicator Cluster Survey (MICS), 2025, a nationally representative household survey implemented by the Bangladesh Bureau of Statistics (BBS) with technical support from UNICEF (17). The survey employed a two-stage stratified cluster sampling design. Urban and rural areas within each of Bangladesh’s 64 districts (along with Dhaka North, Dhaka South, and Chattogram City Corporations) formed the main sampling strata. Within each stratum, census enumeration areas (clusters) were selected systematically with probability proportional to size, followed by a systematic sample of 20 households per cluster. A total of 3,149 clusters and 62,980 households were selected nationally. Details regarding the sampling process can be found in the Bangladesh MICS 2025 survey report (17).

### Study population

For the present analysis, we used the women’s dataset from MICS 2025 Bangladesh. The survey included 64,405 women and 2,711 pregnant women aged 15-49 years were identified during the household survey and selected for blood collection. All identified pregnant women in a household were approached for blood collection. Venous blood samples were collected from 1,940 women who provided consent, of whom 1,934 had valid laboratory results. After excluding observations with missing values on the outcome or covariates of interest (blood lead category, education, arsenic, healthcare decision-making, and pregnancy trimester) under a complete-case approach, 1,928 women constituted the final analytic sample.

### Blood specimen collection

Blood was collected in household settings by 43 dedicated, professionally trained phlebotomists, who used aseptic precautions and sterile, disposable syringes and needles. A 2 mL venous blood specimens were obtained from each participant and transported to the designated reference laboratory in Dhaka: the Immunobiology, Nutrition and Toxicology Laboratory at icddr,b, where heavy metal concentrations were measured using validated methods.

### Heavy metal analysis by ICP-MS

Heavy metals (lead, cadmium, mercury, arsenic) were quantified using an inductively coupled plasma mass spectrometer (ICP-MS; NexION 2200, PerkinElmer, USA). Blood specimens, including standards and controls, were digested with concentrated nitric acid. After digestion, a diluent (a mixture of methanol, nitric acid, gold standard, and type-1 water) and a mixture of internal standards were added to the samples. Digested blood-based working standards, controls (RECIPE Chemicals + Instruments, GmbH), and diluted digested samples were then run into the ICP-MS under optimized spectroscopic conditions. The concentration of each metal was calculated from the respective standard curve prepared using these standards. Additionally, blanks were analyzed to verify the absence of elemental carryover and to confirm low background levels, while three levels of ClinChek Controls (RECIPE Chemicals + Instruments, GmbH) were used to secure both precision and accuracy of daily laboratory results.

Blood levels of four heavy metals/metalloids: lead (Pb, measured in µg/dL), arsenic (As, measured in µg/L), cadmium (Cd, measured in µg/L), and mercury (Hg, measured in µg/L) were quantified. While the official Survey Findings Report deferred estimates for arsenic, cadmium, and mercury in whole blood due to analytical concerns, as whole blood metal, especially arsenic, concentration may not provide a true indication of body burden, this study utilized raw biomonitoring data to investigate underlying public health associations. Findings for these exposures should therefore be interpreted considering the reported analytical limitations.

The process of participant selection is presented in Figure 1.

**Figure 1:**
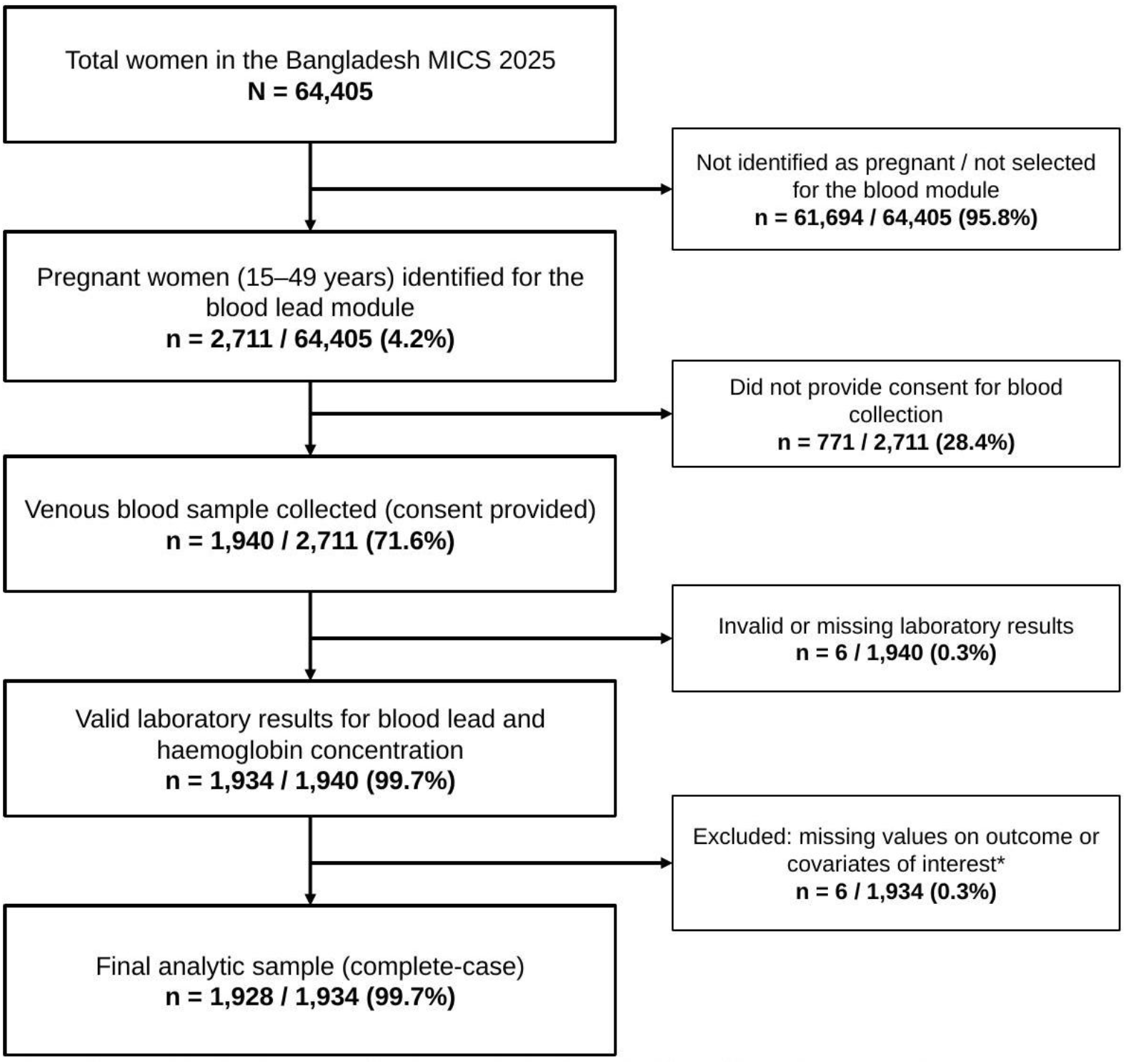
Flow diagram of participant selection, Bangladesh MICS 2025.

### Operational definitions of the independent and the outcome variables

#### Outcome variable

The primary outcome variable was elevated blood lead level (BLL), defined as blood lead concentration ≥5 μg/dL. Blood lead level was dichotomized for the primary analysis. Women with BLL <5 μg/dL was categorized as “absent” and with BLL ≥5 μg/dL as “present”.

#### Independent variables

Blood concentrations of arsenic (μg/L), cadmium (μg/L), and mercury (μg/L) were treated as continuous exposure variables. Sociodemographic and socioeconomic covariates included were age (in years), age at first marriage (in years), area of residence (urban/rural), administrative division (Barishal/Chattogram/Dhaka/Khulna/Mymensingh/Rajshahi/Rangpur/Sylhet), highest level of education completed (up to primary/secondary/post-secondary), wealth index (poorest/poor/middle/rich/richest), participation in healthcare decision-making (no/yes), pregnancy trimester (first/second/third). Observations with values recorded as missing, implausible or don’t know were not included in the analysis.

The “wealth index quintile” variable was derived from “wealth index”, a composite measure of household wealth. To construct the “wealth index” for this MICS 2025 survey, principal components analysis was applied to information on the consumer goods ownership, dwelling characteristics, water and sanitation, and other household’s wealth-associated characteristics, to generate weights (factor scores) for each item. Initial factor scores were first calculated for the total sample. Separate factor scores were then calculated for urban and rural household areas. Next, the urban and rural factor scores were regressed on the initial factor scores to get the combined, final factor scores for the total sample. This was carried out to mitigate urban bias in the wealth index values. Each household in the total sample was assigned a wealth score based on its assets and the final factor scores obtained as described above. The survey household population was then ranked by the wealth score of the household they lived, divided into 10 equal portions (deciles), and then finally regrouped into 5 equal groups (quintiles): lowest (poorest), second (poor), middle, fourth (rich) and highest (richest).

The variable “healthcare decision-making” was recoded from a survey-item that asked, “Who makes decisions for health care of you?”. Participants who responded “respondent” or “joint decision” were grouped together as “participates”. Participants who responded “husband” or “other” was considered as “no participation”.

The variable “pregnancy trimester” considered the gestational age at the time of interview. It has three categories: first trimester (gestational age ≤13 weeks), second trimester (14-27 weeks) and third trimester (≥28 weeks).

### Statistical analysis

All analyses accounted for the complex survey design using Stata’s svy suite (STATA 19, StataCorp LLC, College Station, TX), specifying primary sampling unit, sampling strata, and the normalized blood-module sampling weight for pregnant women. Sampling weights were constructed by BBS/UNICEF as the inverse of selection probability adjusted for non-response, then normalized so that the weighted sample size equaled the unweighted sample size at the national level.

Continuous variables were summarized using survey-weighted means and standard deviations (age, age at marriage) or survey-weighted medians and interquartile ranges (metal biomarkers). Categorical variables were summarized as unweighted frequencies with survey-weighted percentages. Survey-weighted logistic regression was used to estimate unadjusted (crude) odds ratios for the association between independent variables and elevated blood lead, followed by a single multiple models including all covariates simultaneously to estimate adjusted odds ratios (aOR). Both models are reported to have 95% confidence intervals (CIs). For the regression analysis, blood as, cd and hg were log2 transformed for each interpretation where a 1-unit change represents 2-fold rise in the blood heavy metal level.

Socioeconomic inequality in elevated blood lead across wealth quintiles was quantified using the Slope Index of Inequality (SII) and Relative Index of Inequality (RII), estimated via survey-weighted regression-based methods using the healthequal package (18). Population attributable fractions (PAF) were estimated to use counterfactual scenarios based on the fully adjusted logistic regression model, for three counterfactual scenarios: shifting the entire population’s residence to rural, education to secondary level and healthcare decision-making to full participation. PAFs and their 95% Cis were reported.

As a sensitivity analysis, adaptive least absolute shrinkage and selection operator (LASSO) logistic regression was used for data-driven variable selection among all 19 candidate covariates, using 10-fold cross-validation to select the penalization parameter (λ); a fixed random seed (4321) was used for reproducibility. Variables retained by the adaptive LASSO were refit in a survey-weighted logistic regression model to obtain interpretable aORs with their 95% CIs and was compared against the prespecified multivariable model for consistency. Statistical significance was defined as p<0.05 for all analyses.

### Ethical approval

The Bangladesh MICS7 survey plan, questionnaires, and protection protocol, including procedures for biological specimen collection, were reviewed and approved by the icddr,b Ethical Review Committee (reference: PR-24113) (17). Verbal informed consent was obtained from all respondents for the household and individual interviews, and separate written informed consent was obtained before venous blood collection for the blood lead level module. This secondary analysis used de-identified survey data.

## Results

Table 1 presents the distribution of the participants using survey-weighted mean (SD), median (IQR), frequency (unweighted) and percentages (survey-weighted). Among the 1928 participants, 6.2% reported having an elevated BLL. The survey-weighted mean (SD) age of the participants was 24.3 (5.9) years and age at first marriage was 17.4 (3.1) years. Most (70.6%) participants were rural residents and had secondary level of education (56.6%). Regarding healthcare decision making, 71.3% reported participating. The median (IQR) of blood cadmium level was higher among women with elevated BLL (0.8 [0.5-1.2] vs 0.7 [0.4-0.9]). However, the median (IQR) of blood mercury level was lower (1.1 [0.8-1.5] vs 1.5 [1.0-2.2]) (Table 1).

**Table 1:**
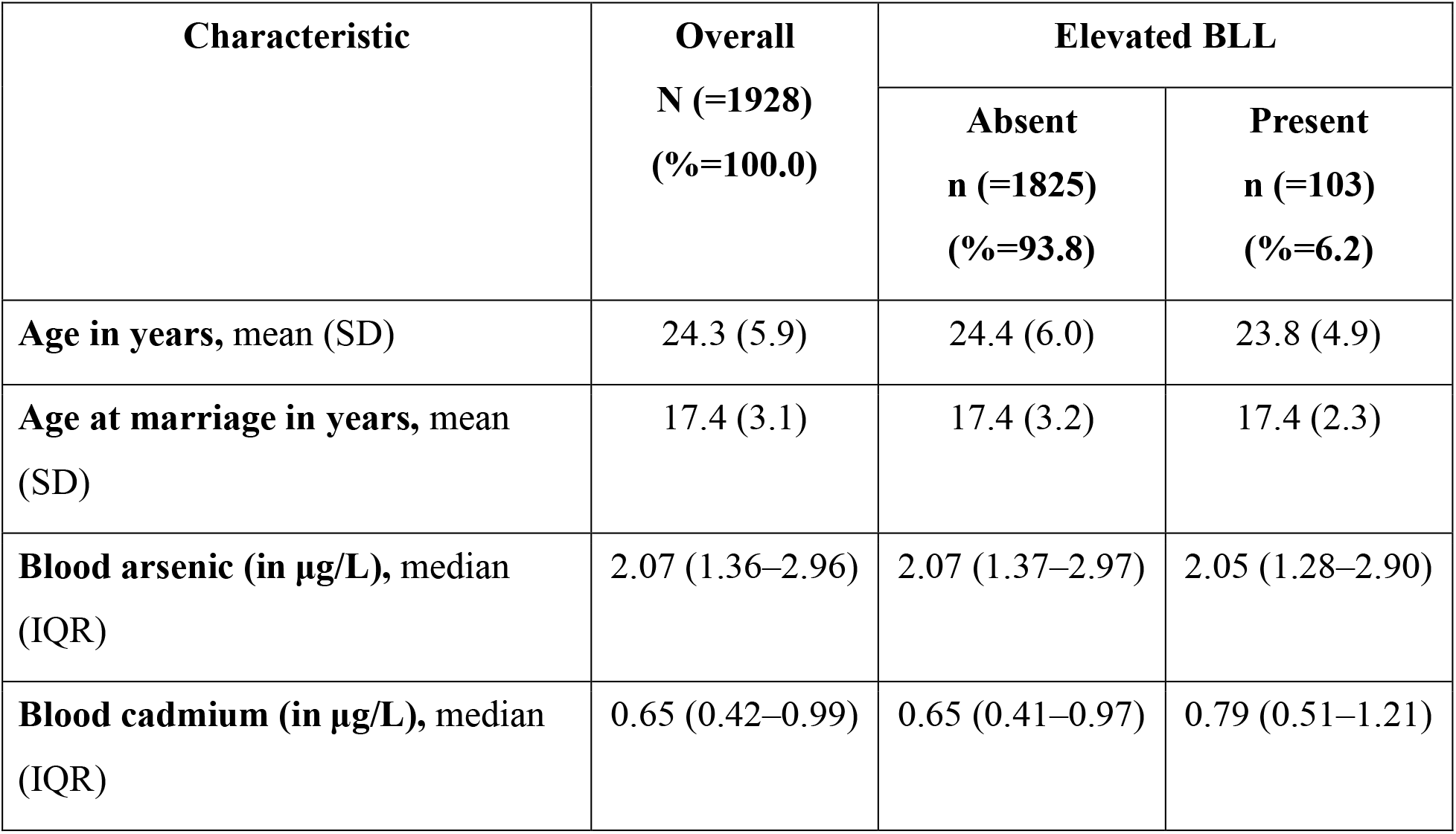
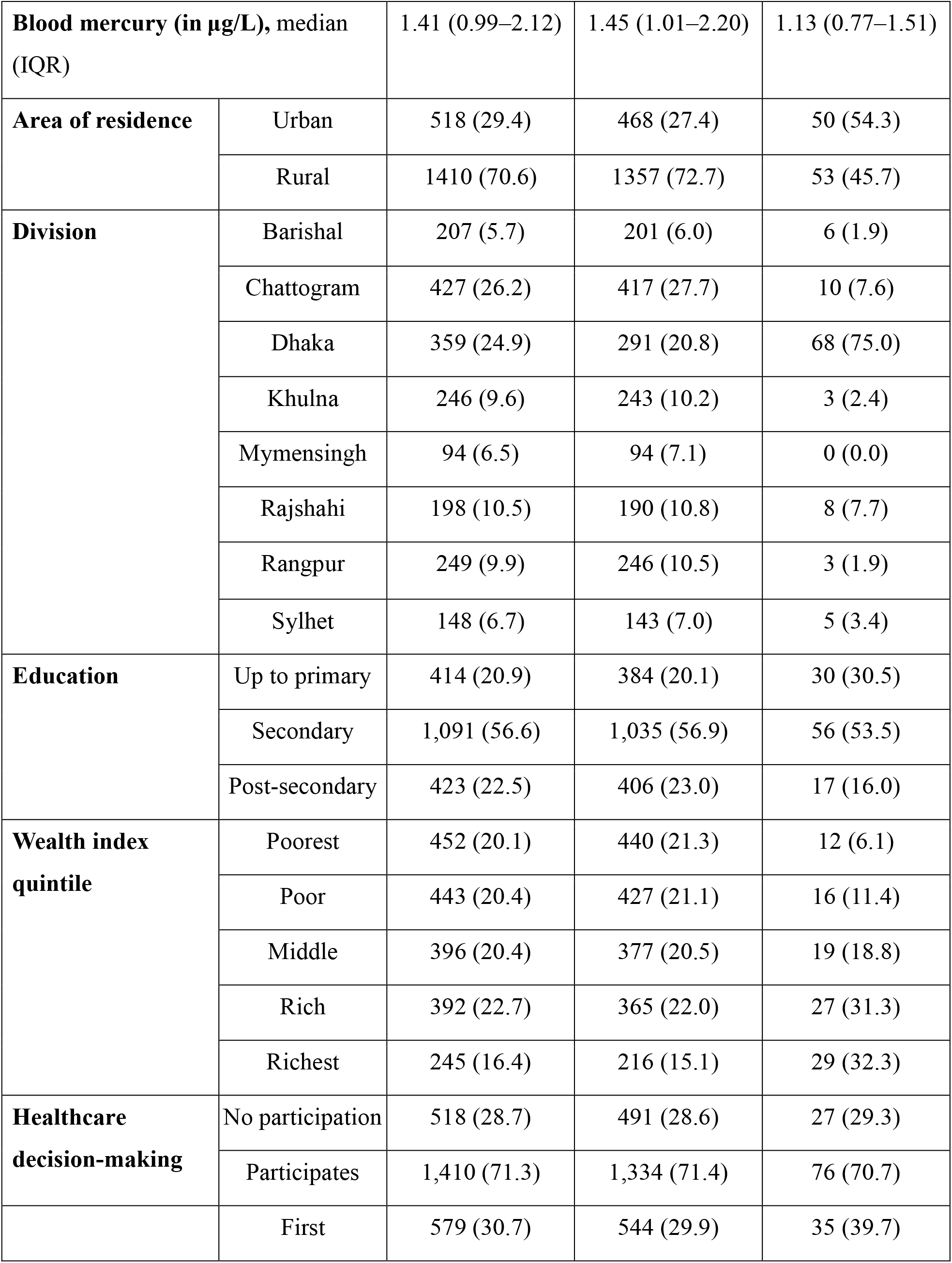
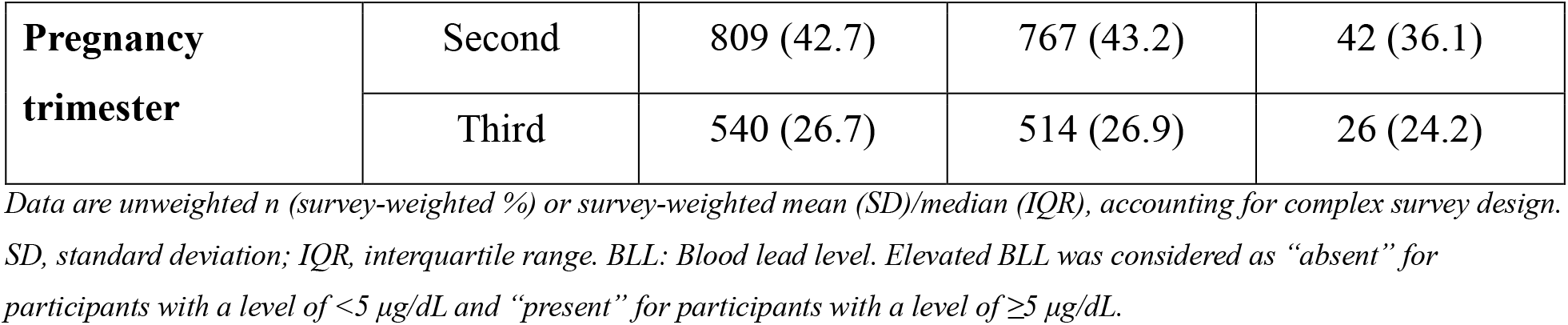
Baseline characteristics of pregnant women, overall and by blood lead level.

In the adjusted model secondary level of education (compared to up to primary), rural residence (compared to urban) and 2-fold rise in blood mercury level was associated with lower odds of elevated BLL (aOR: 0.49; 95% CI: 0.28, 0.85; aOR: 0.55; 95% CI: 0.32, 0.94; and aOR: 0.63; 95% CI: 0.48, 0.83, respectively). However, 2-fold rise in blood cadmium level (aOR: 1.54; 95% CI: 1.21, 1.96), and compared to poorest wealth quintile belonging to poor (aOR: 2.47; 95% CI: 1.04, 5.87), middle (aOR: 4.97; 95% CI: 2.07, 11.92), rich (aOR: 6.14; 95% CI: 2.82, 13.38) and richest quintile was associated with higher odds of elevated BLL (Table 2).

**Table 2:** Unadjusted and adjusted correlates of elevated blood lead level (BLL) among pregnant women.

| Variable | uOR (95% CI) | P value | aOR (95% CI) | P value |
| --- | --- | --- | --- | --- |
| Age in years | 0.98 (0.95–1.02) | 0.398 | 0.97 (0.92–1.01) | 0.154 |
| Age at marriage in years | 1.01 (0.95–1.07) | 0.841 | 1.05 (0.97–1.13) | 0.204 |
| Blood arsenic level* | 0.90 (0.73–1.11) | 0.327 | 1.03 (0.80–1.32) | 0.824 |
| Blood cadmium level* | 1.47 (1.16–1.85) | **0.001** | 1.54 (1.21–1.96) | **<0.001** |
| Blood mercury level* | 0.63 (0.49–0.81) | **<0.001** | 0.63 (0.48–0.83) | **0.001** |
| Residence | Ref. Urban |  |  |  |
| Rural | 0.32 (0.21–0.49) | **<0.001** | 0.55 (0.32–0.94) | **0.030** |
| Education | Ref. Up to primary |  |  |  |
| Secondary | 0.62 (0.38–1.02) | 0.061 | 0.49 (0.28–0.85) | **0.012** |
| Post-secondary | 0.46 (0.23–0.92) | **0.029** | 0.22 (0.11–0.46) | **<0.001** |
| **Wealth index** | Ref. Poorest |  |  |  |
| Poor | 1.86 (0.81–4.30) | 0.145 | 2.47 (1.04–5.87) | **0.040** |
| Middle | 3.18 (1.38–7.34) | **0.007** | 4.97 (2.07–11.92) | **<0.001** |
| Rich | 4.93 (2.36–10.28) | **<0.001** | 6.14 (2.82–13.38) | **<0.001** |
| Richest | 7.42 (3.53–15.60) | **<0.001** | 10.32 (4.41–24.15) | **<0.001** |
| **Healthcare decision** | Ref. No participation |  |  |  |
| Participates | 0.97 (0.59–1.59) | 0.892 | 1.02 (0.60–1.74) | 0.929 |
| **Trimester** | Ref. First trimester |  |  |  |
| Second | 0.63 (0.37–1.07) | 0.089 | 0.65 (0.38–1.12) | 0.121 |
| Third | 0.68 (0.38–1.20) | 0.179 | 0.86 (0.47–1.56) | 0.610 |

OR (95% CI) from separate univariable survey-weighted logistic regression models; aOR (95% CI) from a single multivariable model including all listed variables. Both account for complex survey design. Bold indicates $p < 0.05$ . OR, unadjusted odds ratio; aOR, adjusted odds ratio; CI, confidence interval; \*log<sub>2</sub> transformed, indicated 2-fold rise

Counterfactual scenarios based on the fully adjusted model estimated that shifting the population to secondary-level education was estimated to reduce prevalence by 52.9% (PAF: 52.9, 95% CI:20.6, 72.0) and shifting to rural residence by 24.3% (PAF: 24.3, 95% CI: 01.4, 41.9) (Table 3).

**Table 3:** Population attributable fraction of elevated blood lead under counterfactual exposure scenarios.

| Variable | Counterfactual scenario | PAF (95% CI) |
| --- | --- | --- |
| Residence | All women shifted to Rural | 24.3 (1.4, 41.9) |
| Education | All women shifted to Secondary | 52.9 (20.6, 72.0) |
| Healthcare decision-making | All women shifted to Participates | -0.7 (-17.6, -13.8) |

PAF, population attributable fraction based on the adjusted, survey-weighted logistic regression model.

The SII was 0.16 (95% CI: 0.10, 0.22), indicating a log-scaled difference in predicted prevalence of elevated BLL of approximately 16 percentage points between the richest and poorest ends of the wealth distribution. The RII was 8.59 (95% CI: 4.24, 17.40), indicating that belonging to the richest quintile was associated with 8.59 times higher odds of elevated BLL (Table 4).

**Figure 2:**
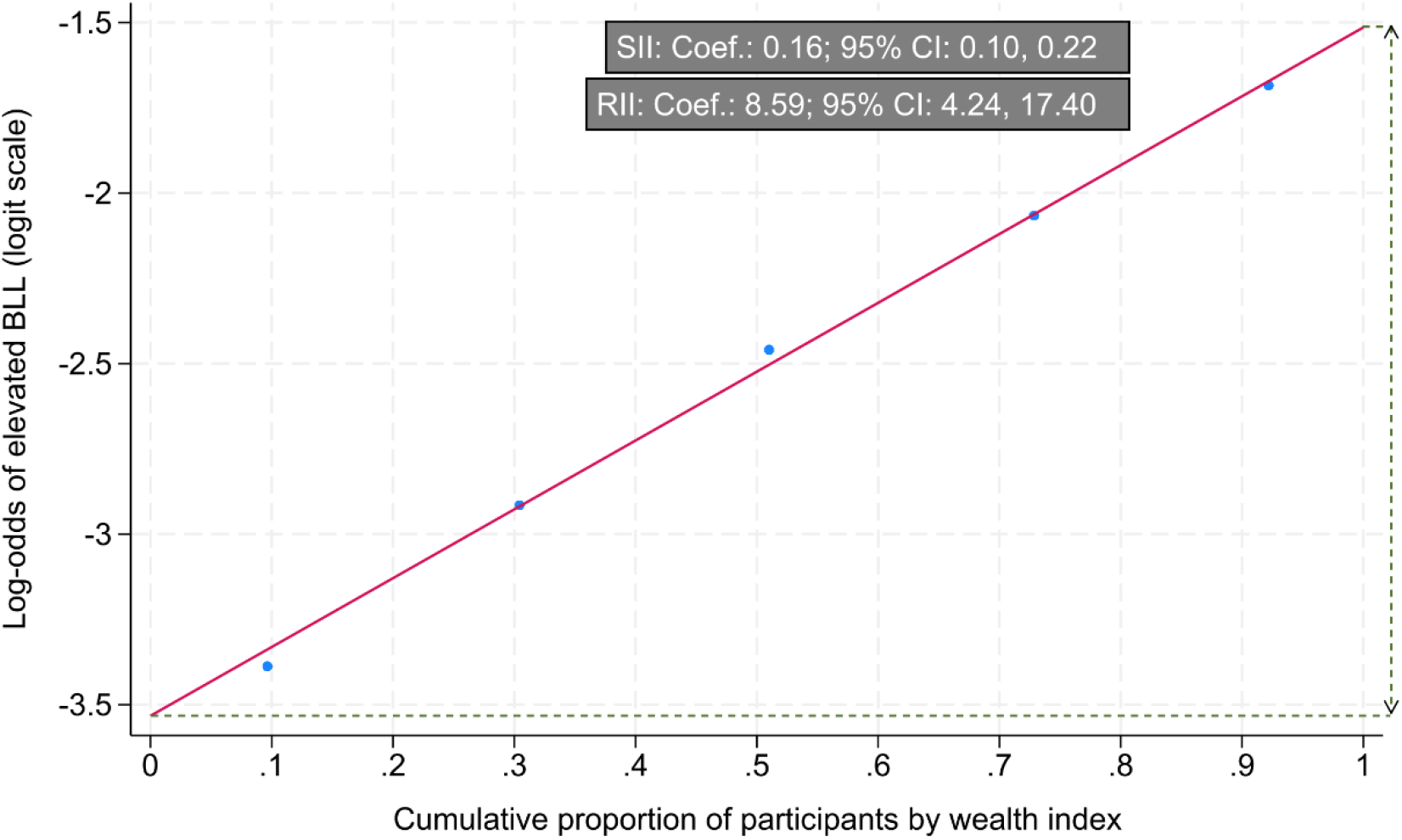
Wealth-Related Inequality in Elevated Blood Lead Level. SII: Slope Index of Inequality; RII: Relative Index of Inequality; CI, confidence interval; Coef.: Coefficient (estimated using logistic regression and are presented on the log-odds scale); BLL: Blood lead level; Coef represents the log-odds (likelihood) of women reporting elevated BLL (postitive values indicate a higher likelihood in richest quintile)

Adaptive LASSO variable selection, performed among 19 candidate covariates using 10-fold cross-validation, retained 9 covariates. The LASSO-selected variables were refit in a survey-weighted logistic regression model, where estimates were consistent in direction and magnitude with the fully adjusted model (supplementary table1).

**Supplementary Table 1:**
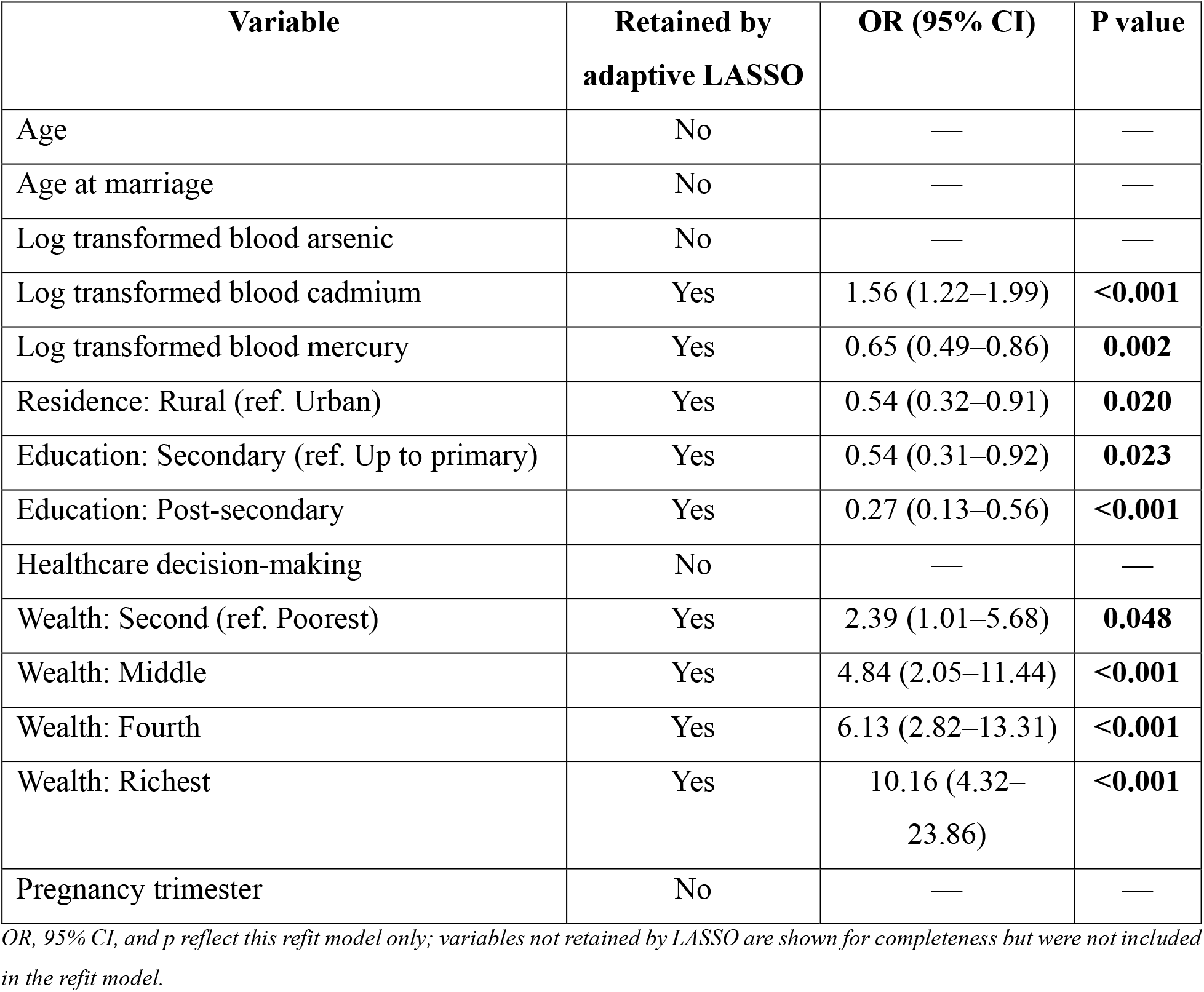
Adaptive LASSO variable selection and refit survey-weighted logistic regression for elevated blood lead level.

| Variable | Retained by adaptive LASSO | OR (95% CI) | P value |
| --- | --- | --- | --- |
| Age | No | — | — |
| Age at marriage | No | — | — |
| Log transformed blood arsenic | No | — | — |
| Log transformed blood cadmium | Yes | 1.56 (1.22–1.99) | <0.001 |
| Log transformed blood mercury | Yes | 0.65 (0.49–0.86) | 0.002 |
| Residence: Rural (ref. Urban) | Yes | 0.54 (0.32–0.91) | 0.020 |
| Education: Secondary (ref. Up to primary) | Yes | 0.54 (0.31–0.92) | 0.023 |
| Education: Post-secondary | Yes | 0.27 (0.13–0.56) | <0.001 |
| Healthcare decision-making | No | — | — |
| Wealth: Second (ref. Poorest) | Yes | 2.39 (1.01–5.68) | 0.048 |
| Wealth: Middle | Yes | 4.84 (2.05–11.44) | <0.001 |
| Wealth: Fourth | Yes | 6.13 (2.82–13.31) | <0.001 |
| Wealth: Richest | Yes | 10.16 (4.32–23.86) | <0.001 |
| Pregnancy trimester | No | — | — |

OR, 95% CI, and p reflect this refit model only; variables not retained by LASSO are shown for completeness but were not included in the refit model.

## Discussion

In this nationally representative analysis of 1,928 pregnant women in Bangladesh, 6.2% had a BLL at or above the WHO action threshold of 5 μg/dL. Elevated BLL was independently associated with higher blood cadmium, while higher blood mercury, rural residence and secondary or higher education were associated with lower odds. A positive wealth gradient was observed, with women in the richest quintile having over ten times the odds of elevated BLL of those in the poorest quintile, a pattern that was mirrored in a pro-rich SII and RII and reproduced by an independent, data-driven variable selection procedure.

The 6.2% prevalence found in this analytic subsample is somewhat lower than the 7.5% reported for pregnant women in the preliminary MICS 2025 national report (17), a difference that most plausibly reflects our restriction to women with complete data on the outcome and all covariates of interest, together with sampling variability once the survey design and weights are accounted for. To our knowledge, MICS 2025 is the first nationally representative survey to measure BLL in pregnant women in Bangladesh, and no directly comparable national baseline therefore exists. The only earlier Bangladeshi estimate in pregnant women came from a purposively sampled rural case-control study of approximately 400 women that specifically enriched for women with extreme BLLs (14); because of that design, its estimate is not comparable to a population-based prevalence and cannot be used to infer a temporal trend. We therefore refrain from drawing any conclusion about changes in population BLL over time. What the present findings do establish is that elevated BLL remains a substantial public health problem in this population, affecting roughly 6% of pregnant women nationally, with exposure patterned by geography, education, and wealth. A government-led campaign against lead-adulterated turmeric and other food-related sources has been implemented in Bangladesh over the past decade. Whether these efforts have reduced population BLL cannot be assessed here, because no comparable national measurement in pregnant women predates the present survey.

The positive association between blood cadmium and elevated BLL is consistent with the two metals sharing common environmental and dietary sources, including contaminated rice, vegetables and tobacco or biomass smoke, and with evidence that cadmium and lead act through overlapping toxicokinetic pathways, including competition for shared transport and binding proteins in the gut and placenta (13, 19). As cadmium has itself has been independently associated with lower birth weight and hypertensive disorders of pregnancy (13, 20), women with elevated levels of both metals may face compounded risk. Subsequent interventions that target a single metal in isolation may only be partially effective for this group.

The inverse association between blood mercury and elevated BLL was unexpected, since mercury is not generally considered biologically protective against lead toxicity, and this finding should be interpreted cautiously. It most likely reflects the fact that mercury and lead in this population arise from largely distinct exposure pathways – mercury predominantly through marine fish consumption and lead through food, dust, cosmetics and occupational contact – so that women whose diet or environment favors one type of exposure may systematically differ, in ways not fully captured by the covariates available in this analysis, from women exposed predominantly to the other (21, 22). Residual confounding by unmeasured dietary or occupational patterns cannot be excluded, and this association merits dedicated source identification investigation rather than mechanistic interpretation.

The strong, positive wealth gradient in elevated BLL is arguably the most impactful finding of this analysis and directly mirrors the pattern reported in the preliminary national MICS report, in which the richest quintile carried the highest burden of childhood and maternal lead exposure (17). This pattern departs from the socioeconomic gradient typically seen for maternal and child health outcomes in Bangladesh, where poorer households usually bear a disproportionate burden of disease, and instead points to exposure sources that track with, rather than against, material affluence. Plausible contributors include greater urban residence among wealthier households and closer proximity to traffic, industrial emissions and informal lead-acid battery recycling operations that are concentrated around cities such as Dhaka (15); greater use of imported cosmetics, ceramics, metal cookware or traditional remedies that can contain lead; and home renovation or construction activity involving lead-based paints or fittings (23). Because these exposures are consumer- and environmental-related rather than markers of poverty, thus conventional poverty-targeted health interventions aimed at poverty are unlikely to reach the women most affected. Additionally, the substantial PAF observed for education (52.9%) suggests that health-literacy-oriented interventions, rather than income-transfer approaches alone, may represent a more promising entry point.

The protective association seen for rural residence and higher education, together with the accompanying PAF estimates, supports the same explanation: urban residence and lower educational attainment may each independently correspond to different dimensions of exposure, ranging between proximity to industrial hotspots and differences in awareness of specific contamination sources such as adulterated spices, leaded cookware or unsafe battery disposal (23). The level of wealth-related inequality quantified here, an RII of 8.59 and an SII representing an absolute gap roughly 16-percentage-point in predicted prevalence between the wealthiest and poorest women, is large relative to what is typically reported for other maternal and child health indicators across low- and middle-income countries (24). This underscores that lead exposure in Bangladesh is not simply a disease of poverty but a distinct equity problem requiring its own targeting logic.

The fact that the adaptive LASSO procedure, applied without any priori model specification, retained nine of the same covariates as the prespecified multivariable model and produced estimates consistent in direction and magnitude increases confidence that these associations are not simply artefacts of variable selection or model specification.

Together, these findings have direct implications for antenatal care and public health policy in Bangladesh. WHO guideline for clinical management of lead exposure recommends that any pregnant woman with a BLL ≥5 μg/dL should have her exposure sources investigated and receive counselling on exposure reduction and nutritional support, including sufficient calcium intake to limit skeletal lead mobilization (4, 9). Integrating blood lead screening within routine antenatal care, at least in high-burden divisions such as Dhaka, and combining this with household-level source investigation rather than an income-based targeting strategy, would align with the exposure pattern identified here. More broadly, as childhood lead exposure in Bangladesh is estimated to cost billions of dollars annually in lost economic productivity (25), reducing lead exposure during pregnancy, when a substantial portion of lifelong neurodevelopmental risk to the next generation is established, represents a high-value investment.

This study has a few limitations. First, its cross-sectional design doesn’t allow causal inference, and reverse causation or residual confounding cannot be nullified for any of the reported associations. Second, restricting the analytic sample to women with complete data on both the outcome and all covariates, although necessary for the regression approach employed, could introduce selection bias if data completeness was systematically related to BLL; the modest difference between our subsample prevalence and the national estimate suggests this effect was likely small but cannot entirely be ruled out. Third, BLL was measured at a single time point and does not reflect trimester-specific dynamics in maternal lead mobilization, nor does it distinguish current environmental exposure from lead released from long-term skeletal stores. Fourth, detailed information on specific exposure sources, such as occupational lead contact, use of cosmetics or traditional remedies, dietary patterns, and iodine or calcium intake, was not available and could not be adjusted for, leaving open the precise mechanisms underlying the wealth and mercury associations reported here. Fifth, the number of women with elevated BLL (n = 103) was modest, limiting statistical power for subgroup analyses and widening the confidence intervals around several estimates, most visibly for the wealth index categories. Finally, as with any PAF estimated from an observational model, our counterfactual estimates assume that the fitted model correctly captures the exposure–outcome relationship and should be interpreted as indicative of the scale of potential impact rather than as precise causal effects.

In conclusion, elevated blood lead level affected an estimated 1 in 16 pregnant women in this nationally representative Bangladeshi sample and was compounded by cadmium co-exposure. Contrary to the pattern seen for most maternal and child health indicators, this burden was concentrated among wealthier, urban and less-educated women, pointing to consumer and environmental exposure pathways rather than material deprivation as the principal drivers. These findings support the integration of blood lead screening into antenatal care in high-burden settings, targeted investigation of household and consumer sources of lead among wealthier and urban populations and continued national surveillance to track progress as Bangladesh works towards its stated goal of a lead-free future.

### Patient and Public Involvement

Patients and/or the public were not involved in the design, or conduct, or reporting, or dissemination plans of this research.

### Patient Consent for Publication

Not applicable.

## Data Availability Statement

The dataset used in this study are publicly available from the respective data providers. The 2025 MICS datasets are available through the UNICEF MICS data repository, subject to the applicable data access requirements (https://mics.unicef.org/surveys). The data used in this study were obtained through the respective data access procedures.

